# Efficacy and safety of chloroquine or hydroxychloroquine in moderate type of COVID-19: a prospective open-label randomized controlled study

**DOI:** 10.1101/2020.06.19.20136093

**Authors:** Lan Chen, Zhen-Yu Zhang, Jian-Guo Fu, Zhi-Peng Feng, Su-Zhen Zhang, Qiu-Ying Han, Xiao-Bin Zhang, Xiong Xiao, Hui-Min Chen, Li-Long Liu, Xian-Li Chen, Yu-Pei Lan, De-Jin Zhong, Lan Hu, Jun-Hui Wang, Xing-Hua Yu, Dan-Yang She, Yong-Hong Zhu, Zhen-Yu Yin

## Abstract

The outbreak of novel coronavirus disease 2019 (COVID-19) has become a pandemic. Drug repurposing may represent a rapid way to fill the urgent need for effective treatment. We evaluated the clinical utility of chloroquine and hydroxychloroquine in treating COVID-19.

Forty-eight patients with moderate COVID-19 were randomized to oral treatment with chloroquine (1000 mg QD on Day 1, then 500 mg QD for 9 days; n=18), hydroxychloroquine (200 mg BID for 10 days; n=18), or control treatment (n=12).

Adverse events were mild, except for one case of Grade 2 ALT elevation. Adverse events were more commonly observed in the chloroquine group (44.44%) and the hydroxychloroquine group (50.00%) than in the control group (16.67%). The chloroquine group achieved shorter time to clinical recovery (TTCR) than the control group (P=0.019). There was a trend toward reduced TTCR in the hydroxychloroquine group (P=0.049). The time to reach viral RNA negativity was significantly faster in the chloroquine group and the hydroxychloroquine group than in the control group (P=0.006 and P=0.010, respectively). The median numbers of days to reach RNA negativity in the chloroquine, hydroxychloroquine, and control groups was 2.5 (IQR: 2.0-3.8) days, 2.0 (IQR: 2.0-3.5) days, and 7.0 (IQR: 3.0-10.0) days, respectively. The chloroquine and hydroxychloroquine groups also showed trends toward improvement in the duration of hospitalization and findings on lung computerized tomography (CT). This study provides evidence that (hydroxy)chloroquine may be used effectively in treating moderate COVID-19 and supports larger trials.

## Introduction

The rapid spread of COVID-19 caused by severe acute respiratory syndrome coronavirus-2 (SARS-CoV-2) has posed an enormous challenge to public healthcare across the globe [1-3]. The World Health Organization (WHO) declared the outbreak of COVID-19 as a pandemic on March 11, 2020 [4]. By April 13, it had affected more than 1.7 million individuals.

As SARS-CoV-2 is a newly identified pathogen, no target-specific antiviral drugs or vaccines are currently available. The adoption of a crisis intervention approach will be required. Additionally, if there is scientific merit, the repurposing of existing drugs with mechanistic potential may provide a quick yet cost-effective treatment paradigm.

Chloroquine and hydroxychloroquine have gained broad attention as possible treatment options for COVID-19, based on three main rationales. First, chloroquine has demonstrated broad-spectrum antiviral activity against viruses such as human immunodeficiency virus, Zika virus, dengue virus, and coronavirus OC43 [5]. After the outbreak of COVID-19 in Wuhan, China, Wang et al. screened existing antiviral drugs as potential treatments for COVID-19. The results showed that chloroquine phosphate has potent antiviral activity against SARS-CoV-2 in vitro [6]. The drug also has a high selection index (SI) at low concentrations [7,8]. Secondly, concentrations of chloroquine in the lungs may be hundreds of times higher than those in plasma [9]. This unique distribution pattern is conducive to the treatment of COVID-19. We analyzed the reported EC_90_ of chloroquine phosphate against SARS-CoV-2 in Vero E6 cells reported by Wang et al. [6] and hypothesized that this reported EC_90_ is potentially achievable in humans when the licensed dose of chloroquine phosphate is administered. Third, chloroquine and hydroxychloroquine are immune modulators that affect multi-aspects of the immune response [10]. Both compounds block the production of inflammatory cytokines such as interferon, tumor necrosis factor, interleukins, and B-cell activation factor [5,10]. Additionally, chloroquine and hydroxychloroquine inhibit the interaction between Toll-like receptors 7/8/9 and nucleic acid ligands, thus blocking the activation of innate immunity [8]. These effects may also suppress the formation of cytokine storms in COVID-19 pathogenesis [5,11]. Finally, chloroquine may down-regulate the expression of coinhibitory molecules expressed by plasmacytoid dendritic cells, such as IDO and PD-L1 that suppress T-cell activation [12]. Chloroquine thus positively regulates certain aspects of immune function, in addition to its broad immunosuppressive effects.

Based on these rationales, we investigated the therapeutic utility of chloroquine in treating COVID-19. Hydroxychloroquine is a derivative of chloroquine with an improved safety profile. We designed this randomized, open-label, 3-arm, prospective study to compare the effects of chloroquine or hydroxychloroquine with the standard of care for treating COVID-19. The study was conducted at a hospital in Wuhan. During the study period, the China National Health Commission published the 6^th^/7^th^ editions of “Diagnosis and Treatment Protocol for Novel Coronavirus Pneumonia”, both of which recommend the use of chloroquine [13,14].

This single-center, prospective study was to evaluate the effects of chloroquine phosphate or hydroxychloroquine sulfate in treating COVID-19. In the original study design, we had planned to recruit 100 subjects with confirmed mild/moderate types of COVID-19. However, the city of Wuhan rapidly controlled the epidemic, and we had to complete the study on March 30, 2020, after enrolling only 67 subjects with mild/moderate COVID-19. Here we report our data on 48 patients with moderate type of COVID-19. We provide evidence that patients treated with either chloroquine or hydroxychloroquine showed a positive trend toward faster clinical recovery, more rapid return to viral RNA negativity, and shorter hospitalization. Both molecules had manageable safety profiles at dosage regimens similar to those used to treat extraintestinal amebiasis or rheumatoid arthritis. We hope that the data provided will serve as reference and justification for the further investigation and use of (hydroxy)chloroquine by healthcare professionals across the globe who are fighting COVID-19.

## Methods

### Study design and participation

From Feb 18 to Mar 30, 2020, we conducted a prospective, open-label, randomized and controlled study (Clinical Trial Registration Number: ChiCTR2000030054) at Ward E3-9, Optical Valley Campus of Tongji Hospital, Wuhan, Hubei Province, China. Our study team managed Ward E3-9 from Zhongshan Hospital, Xiamen University, Xiamen, Fujian Province. The institution’s ethics committee approved this study. Prior to screening, all enrolled patients signed their written informed consent.

Eligible patients were 18 to 75 years of age, inhabitants of the Wuhan area, and diagnosed with mild or moderate types of COVID-19 based on the Chinese Diagnosis and Treatment Protocol for Novel Coronavirus Pneumonia (5^th^ −7^th^ Editions) [13-15]. Patients had positive results on reverse transcription polymerase chain reaction (RT-PCR) test for SARS-CoV-2 or lung changes characteristic of COVID-19 on computerized tomography (CT) scan of the chest. At admission, all patients had SaO_2_ (oxygen saturation) > 93% at room temperature. Fully detailed inclusion/exclusion criteria are provided in the robust synopsis (See supplementary information.). In the paragraphs below, we present results from patients with moderate COVID-19. Moderate COVID-19 is defined as: 1. cases meeting the diagnostic criteria for COVID-19; 2. pneumonia confirmed by chest CT scan; 3. absence of severe hypoxia or dyspnea (SaO_2_>93%, PaO_2_/FiO_2_>300 mmHg, respiratory rate <30/min) [13-15]. The trial was designed to assess the efficacy and safety/tolerability of chloroquine and hydroxychloroquine, compared to the standard of care.

### Randomization and masking

This study was an open-label study. Eligible patients were randomized at a 2:2:1 ratio to one of three treatment groups using a computer-generated randomization number. Each patient received one of the following treatment options: oral chloroquine phosphate/standard treatment, hydroxychloroquine sulphate/standard treatment and standard treatment only.

### Procedures

After randomization, all patients received standard treatment according to the Chinese Diagnosis and Treatment Protocol for Novel Coronavirus Pneumonia (5^th^ Edition) [15]. In the chloroquine group, patients received standard treatment and chloroquine phosphate (Guangdong Zhongsheng Pharmaceutical Co., Ltd., Guangzhou, China) orally at 1000 mg QD for the first day, then 500 mg QD for additional 9 days. In the hydroxychloroquine group, patients received standard treatment plus oral hydroxychloroquine sulphate (Shanghai Pharmaceutical Co., Ltd., Shanghai, China) at 200 mg BID for 10 days.

The study period was 28 days or prior to hospital discharge. Patients were assessed following the robust synopsis (available on-line). Vital signs and parameters reflecting clinical recovery (e.g. body temperature, respiratory rate, SaO2, cough) and safety evaluation were performed daily. Laboratory exams (e.g. complete blood count, blood chemistry, C-reactive protein (CRP), procalcitonin) were performed. Patients were also tested with cardiac enzyme panel (troponin, myoglobin and creatine kinase-MB (CK-MB)) at the time of screening and on Day 7. If patients had abnormal levels of cardiac enzyme(s) at baseline, one additional test was done on Day 3 or 4. All patients received a quick bedside cardiac monitoring on Day 3 or 4. If any abnormality was observed, the patient would receive a comprehensive Electrocardiography (ECG) test. SARS-CoV-2 RT-PCR and chest CT were performed during the study and/or prior to discharge.

Additionally exploratory biomarkers were measured: cytokines (IL-1β, IL-2R, IL-6, Il-8, IL-10, and TNFα) and SAR-CoV-2-specific IgG/IgM antibody.

### Outcomes

The primary outcome measurement for this study was time to clinical recovery (TTCR), defined as the number of days from randomization to clinical recovery. Maximum TTCR was 28 days. Patients were considered to have achieved clinical recovery when they had met all of the following criteria for at least 48 hours: 1. axillary body temperature ≤36.9°C or oral body temperature ≤37.2°C; 2. complete relief of all symptoms other than cough; 3. cough graded as mild or absent on a patient-reported scale of severe, moderate, mild, absent.

Secondary outcome measurements included (up to 28 days): 1. time to SARS-CoV-2 RNA negativity, defined as the time from randomization to the day of the first test in two consecutive tests of nasopharyngeal or oropharyngeal swabs with negative SARS-CoV-2 RNA; 2. length of hospital stay, defined as the number of days from randomization to discharge; 3. Changes on chest CT scan; 4. duration (days) of supplemental oxygenation; 5. frequency of adverse events; 6. clinical status; 7. all-cause mortality; 8. duration (days) of supplemental oxygenation; 9. vital signs; 10. results of laboratory testing.

We used a semi-quantitative scoring system similar to the method reported by Pan et al. to estimate the pulmonary manifestations on CT scan [16]. The five lobes of the lungs were assigned a total score of 20 points, based on the volume of tissue affected. The score was given based on the overall affected areas of each side of the lung. Every 10% of affected areas from each lung can be recorded 1 point. Changes in cytokine levels and COVID-19-specific IgG/IgM levels were measured during the first week of hospitalization.

### Statistical analysis

Measurement data were tested for normality. Normally distributed measurement data were described as mean ± standard deviation. Non-normally distributed measurement data were shown as median (interquartile range). Statistical analysis of the primary endpoint was conducted using a Kaplan-Meier curve and Logrank testing. Primary comparisons included both chloroquine vs. control and hydroxychloroquine vs. control. Taking an overall alpha=0.05 as the test standard, we deemed difference with P<0.025 as statistically significant. All other statistical analyses were considered as exploratory. SPSS v17.0 software was used for statistical analysis.

## Results

From Feb 18, 2020 to Mar 30, 2020, we evaluated 94 hospitalized patients with mild/moderate COVID-19. Sixty-seven patients were enrolled into the study, which included a chloroquine group (n=25), a hydroxychloroquine group (n=28), and a control group (n=14). Patients were randomly assigned to treatment groups at a ratio of 2:2:1 (Figure 1). Seven patients in the chloroquine arm, 6 patients in the hydroxychloroquine arm, and 2 patients in the control arm were excluded from analysis because COVID-19 manifested as mild symptoms only. There were an additional 4 patients in the hydroxychloroquine arm who were misdiagnosed with COVID-19 and therefore excluded. We thus reported on 48 cases of moderate COVID-19: 18 in the chloroquine arm (2 were asymptomatic), 18 in the hydroxychloroquine arm (3 were asymptomatic), and 12 in the control arm (Figure 1). The chloroquine arm had an average age of 45.22 ± 13.66 years, including 7 males (38.89%). The hydroxychloroquine arm had an average age of 45.67±14.37 years and included 8 males (44.44%). The control arm had an average age of 51.33 ± 15.36 years with 7 males (58.33%). The three treatment groups were balanced in terms of age, gender, body mass index (BMI), and pre-existing conditions (Table 1).

**Table 1.** Demographic characteristics and pre-existing conditions in patients with moderate COVID-19.

|  | **Chloroquine**<br>**( n=18 )** | **Hydroxychloroquine**<br>**(n=18)** | **Control**<br>**(n=12)** | **P-value** |
| --- | --- | --- | --- | --- |
| **Mean age (years)** | 45.22 ± 13.66 | 45.67 ± 14.37 | 51.33 ± 15.36 | 0.473<sup>a</sup> |
| <40 (years) | 6 | 9 | 4 |  |
| 40-60 (years) | 10 | 6 | 5 | 0.617<sup>b</sup> |
| 60-75 (years) | 2 | 3 | 3 |  |
| **Gender** |  |  |  | 0.572<sup>b</sup> |
| Female | 11 (61.11%) | 10 (55.56%) | 5 (41.67%) |  |
| Male | 7 (38.89%) | 8 (44.44%) | 7 (58.33%) |  |
| **BMI** | 23.19 ± 2.25 | 24.35 ± 3.53 | 23.34 ± 2.60 | 0.442<sup>a</sup> |
| **Preexisting conditions** |  |  |  |  |
| None | 11(61.11%) | 11(61.11%) | 6(50.00%) | 0.796<sup>b</sup> |
| Hypertension | 4 (22.22%) | 3(16.67%) | 1 (8.33%) |  |
| Diabetes | 3 (16.67%) | 3(16.67%) | 3 (25.00%) |  |
| Other* | 2 (11.11%) | 3(16.67%) | 3 (25.00%) |  |

\*: Pre-existing conditions included cardiovascular disease, malignant tumor, chronic obstructive pulmonary disease, chronic kidney disease, anemia, gout, and hypothyroidism.
a: Analysis of variance (ANOVA)
b: $\chi^2$ test

**Figure 1.**
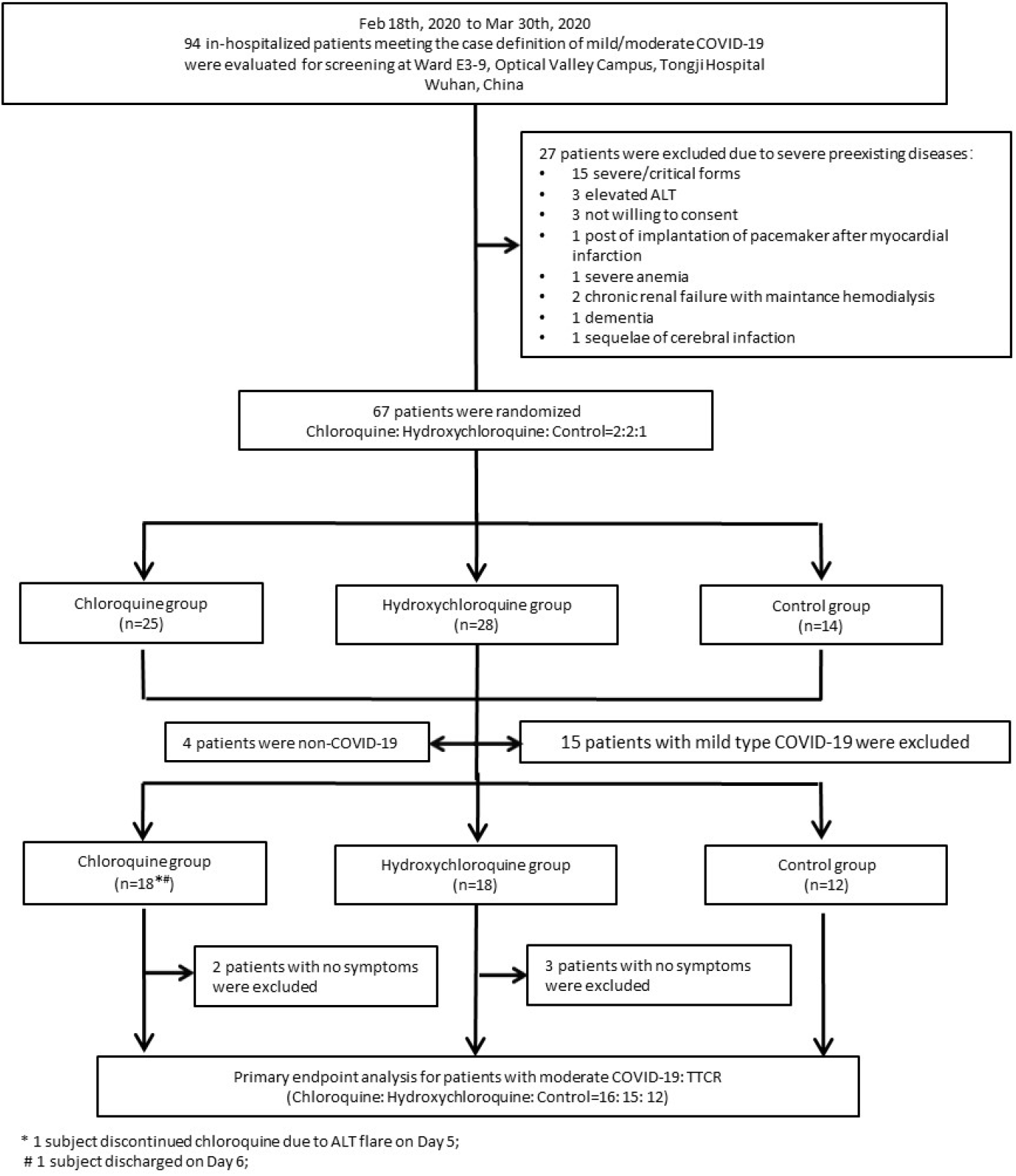
Flow-chart of the study * 1 subject discontinued chloroquine due to ALT flare on Day 5 # 1 subject discharged on Day 6

At baseline, all groups were similar in terms of the frequency of pyrexia, peripheral SaO_2_, requirement for oxygen therapy at admission, and pulmonary CT score (Supplementary Table 1). Additionally, there were no significant differences among groups in the results on baseline laboratory tests, including levels of cytokines (IL-1, IL-2R, IL-6, IL-8, IL-10, TNFα), liver/cardiac enzymes, and SARS-CoV-2-specific IgG/IgM (Supplementary Table 1).

None of the patients included in the study progressed from moderate COVID-19 to severe or critical forms of the disease. There was no death.

Chloroquine and hydroxychloroquine were generally safe and well tolerated. Adverse events (including abnormal laboratory results) were all mild, except for one instance of Grade 2 ALT elevation (CTCAE grading). These mild adverse events were more common in the chloroquine group (44.44%) and the hydroxychloroquine group (50.00%) than in the control group (16.67%) (Table 2). The incidence of diarrhea was higher in the chloroquine group and hydroxychloroquine group (16.67% and 16.67%, respectively) than in the control group (0%). Two patients had ALT/AST elevation. One patient in the chloroquine group had Grade 1 (CTCAE grading) ALT and AST elevation on Day 5 and chloroquine treatment was discontinued. On Day 15, this patient had an ALT level of 57 U/L and a normal AST level. The other patient in the hydroxychloroquine group had an ALT level of 97 U/L and an AST level of 52 U/L prior to randomization. On Day 5, this patient developed Grade 2 ALT elevation and Grade 1 AST elevation. On Day 8, the patient still had Grade 2 ALT elevation and Grade 1 AST elevation. The patient continued to receive hydroxychloroquine until Day 10. On Day 13, the patient’s ALT level decreased to Grade 1; the AST elevation decreased but remained Grade 1.

**Table 2.** Summary of adverse events, n(%).

| Adverse Events | Chloroquine | Hydroxychloroquine | Control |
| --- | --- | --- | --- |
| ALT elevation | 1(5.56) | 1(5.56) | 0(0.00) |
| AST elevation | 1(5.56) | 1(5.56) | 0(0.00) |
| Diarrhea | 3(16.67) | 3(16.67) | 0(0.00) |
| Nausea | 2(11.11) | 3(16.67) | 2(16.67) |
| Fatigue | 0(0.00) | 1(5.56) | 0(0.00) |
| Chest tightness | 1(5.56) | 0(0.00) | 0(0.00) |
| Any adverse events | 8(44.44) | 9 (50.00) | 2(16.67) |

In this study, ECG results obtained at the time of screening were used to exclude patients with prior history of cardiac diseases. We originally planned to include a comprehensive ECG evaluation as part of the study. However, due to the open setting of the ward and our desire to minimize repetitive contact between hospital staff members and patients, we were not able to perform a full ECG assessment for every patient, as originally planned. Nevertheless, a cardiac monitoring for each patient was conducted on Day 3 or 4. No patient had any apparent cardiac abnormality. Patients who had abnormal levels of cardiac enzymes (troponin, myoglobin, and CK-MB) at the time of screening were tested again on Day 3 or 4. In all cases, the results were within normal limits. All patients underwent an additional cardiac enzyme panel on Day 7; no abnormality was observed, in any case (Supplementary Table 2).

We used TTCR as primary endpoint. Kaplan-Meier TTCR curves were generated to compare chloroquine vs. control and hydroxychloroquine vs. control (Figure 2a). Compared with patients receiving standard treatment, patients who received chloroquine treatment had faster TTCR (Logrank (Mantel-Cox) test, P=0.019). The patients who received hydroxychloroquine also showed a trend toward shorter TTCR, compared with the control group (Logrank (Mantel-Cox) test, P=0.049) though it did not reach statistical significance. Additional exploratory Logrank testing (P=0.035) among the three groups revealed the heterogeneity of TTCR responses that were driven by therapeutic benefits of chloroquine and hydroxychloroquine. Finally, median day (IQR) of TTCR for chloroquine group, hydroxychloroquine group, and control group were 5.50 (3.25-7.50) days, 6.00 (3.00-8.00) days, and 7.50 (5.00-16.25) days, respectively. Among the patients in the chloroquine and hydroxychloroquine groups, the longest TTCRs were 10 days and 13 days, respectively. After asymptomatic patients were excluded from statistical analysis, 6.25% (1/16) of those in the chloroquine group and 6.67% (1/15) of those in the hydroxychloroquine group had TTCR ≥10 days. In the control group, the maximum TTCR was 20 days; TTCR was ≥10 days in 41.67% (5/12) of the patients.

**Figure 2a.**
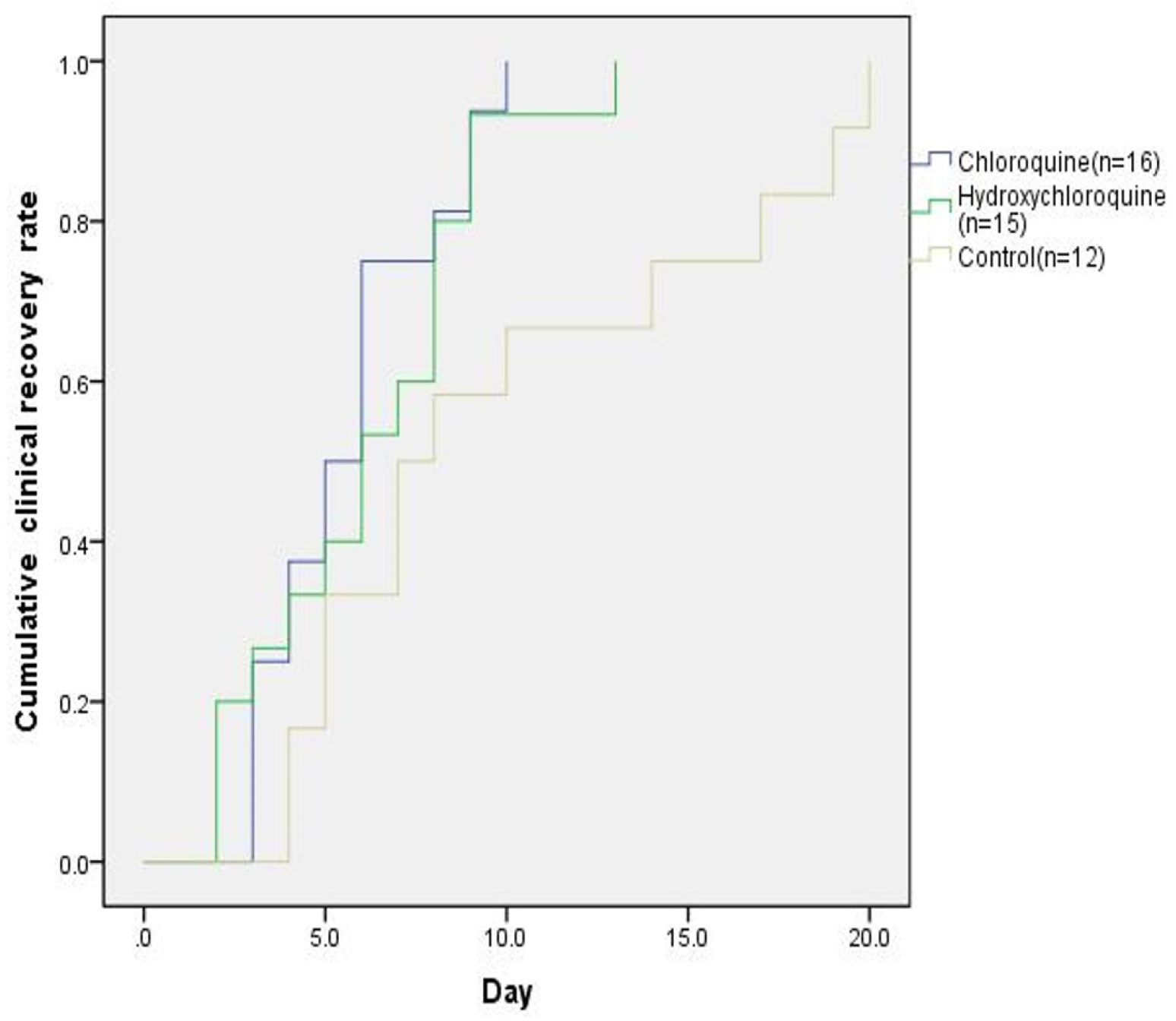
Kaplan-Meier plot analysis for time to clinical recovery (TTCR) in patients with clinical symptoms. Results on the Logrank (Mantel-Cox) test were as follows: chloroquine vs. control, P=0.019; hydroxychloroquine vs. control, P=0.049; comparison among all three groups, P=0.035.

Next, we evaluated time to reach viral negativity by RT-PCR. All patients in the three groups reached viral negativity, as determined by RT-PCR. Compared with the control group [median day: 7.0 (IQR: 3.0-10.0) days], the chloroquine group [median day: 2.5 (IQR: 2.0-3.8) days] and the hydroxychloroquine group [median day: 2.0 (IQR: 2.0-3.5) days] had significant decreases in the number of days required to reach RT-PCR negativity (P=0.006 and P=0.010 by Logrank (Mantel-Cox) test, respectively) (Figure 2b).

We also analyzed days from hospitalization/randomization to discharge (Figure 2c). There was a modest trend toward decrease in the duration of hospitalization for both the chloroquine and hydroxychloroquine groups, consistent with the directional changes in TTCR and time to reach RNA negativity (Figure 2c).

We evaluated chest CT score on Day 7, compared with baseline, and the number of days from randomization to the termination of oxygen therapy (Table 3). The chest CT scores in the chloroquine and hydroxychloroquine groups showed a trend toward improvement while the latter did not show any improvement.

**Figure 2b.**
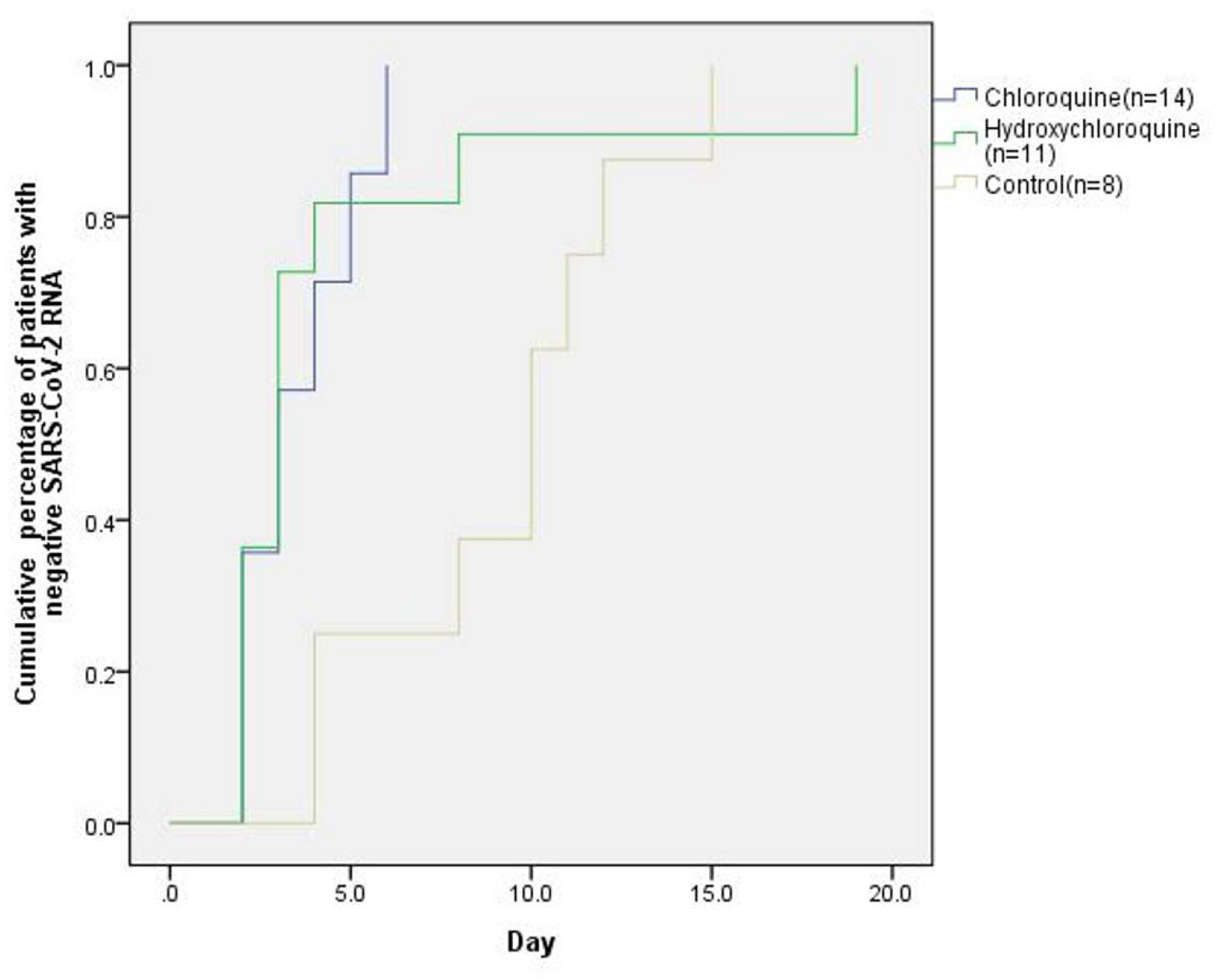
Kaplan-Meier plot analysis for time to SARS-CoV-2 RNA negativity (days from randomization to SARS-CoV-2 RNA negativity) in patients with positive results on SARS-CoV-2 RNA testing at the time of randomization. Results from the Logrank (Mantel-Cox) test were as follows: chloroquine vs. control, P=0.006; hydroxychloroquine vs. control, P=0.010; comparison among all three groups, P=0.017.

**Figure 2c.**
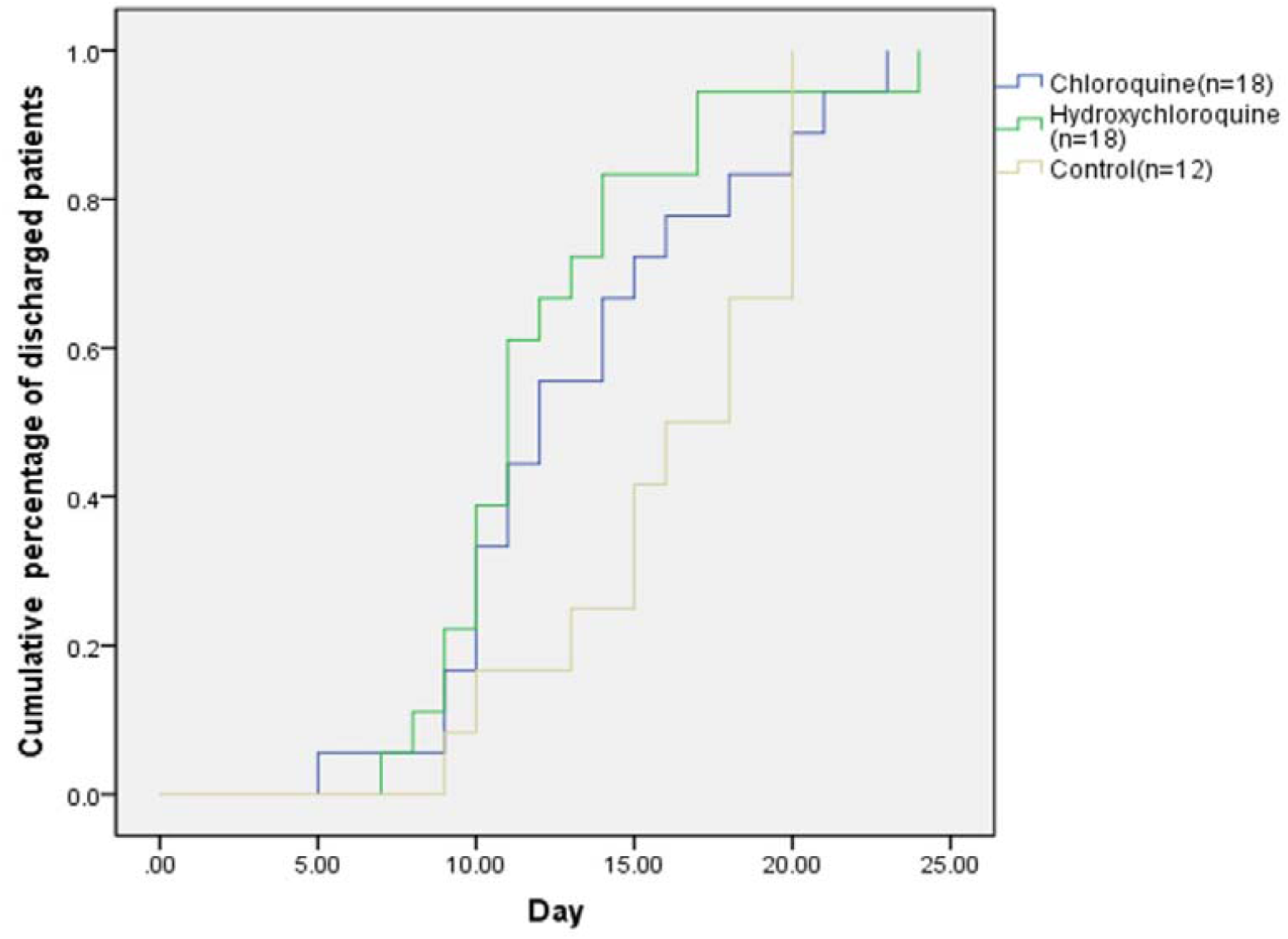
Kaplan-Meier plot analysis for time to discharge (days from randomization to discharge) in patients with moderate COVID-19. Results from the Logrank (Mantel-Cox) test for a comparison among all three groups: P=0.151.

**Table 3.** Other outcomes of additional endpoints among the moderate COVID-19.

|  | Chloroquine |  | Hydroxychloroquine |  | Control |  | P value* |
| --- | --- | --- | --- | --- | --- | --- | --- |
|  | median (IQR) | n | median (IQR) | n | median (IQR) | n |  |
| CT score on the 7th day after randomization | 2.00(0-2.00) # | 17 | 0.00(0-2.00)# | 17 | 2.50(1.50-5.25) | 10 | 0.013 |
| Days from randomization to termination of oxygen therapy | 8.50(0-9.25) | 13 | 7.00(0-9.00) | 12 | 8.00(3.25-14.00) | 10 | 0.277 |

\*: Kruskal-Wallis test
#: $P < 0.05$ , compared with the control group.

Our analyses of cytokines and SARs-CoV-2-specific IgG/IgM changes between Day 1 and Day 7 did not reveal any significant differences (Supplementary Table 2).

## Discussion

We conducted this clinical trial while the study team was in the area of the ongoing COVID-19 epidemic. This study demonstrated that, compared to the control treatment, chloroquine phosphate treatment had significantly improved TTCR in moderate form of COVID-19 (P=0.019). Additionally, chloroquine phosphate treatment accelerated the time to viral RNA negativity, shortened hospital stay, and improved chest CT scan scores. We observed similar trends in the hydroxychloroquine treatment group at the dosage regimen we chose, albeit to a lesser degree. Although comparisons among groups in terms of the time to discharge did not reveal significant difference (P=0.151), the median numbers of days in hospital were lower in the chloroquine and hydroxychloroquine groups than in the control group. As this was a prospective, randomized controlled trial with consistent criteria for enrollment, the three arms were balanced in terms of age, gender, underlying disease, and other baseline characteristics. Our study thus largely avoided issues common to non-randomized trials.

We were inspired to conduct this study by the results of *in vitro* research on SARS-CoV-2 [6]. The robust antiviral activity of chloroquine phosphate against SARS-CoV-2 *in vitro* and the unique pattern of distribution in enriched lung tissue distribution (200 times greater than the plasma concentration) [9]) make it potentially useful for the treatment of COVID-19 [17, 18].

This study generated positive evidence supportive of using (hydroxy)chloroquine to treat moderate forms of COVID-19. However, the data must be interpreted with caution. There were a couple of limitations in the study. First, due to the unexpected onset of this epidemic and the availability of active drugs only, we were not able to conduct a placebo-controlled, double-blinded study, which would have been more objective and largely excluded the possible impact of open-label use on a given physician’s assessment. Second, because the Chinese government implemented a robust intervention approach that effectively controlled the epidemic, the study was terminated before full enrollment (n=40 each for the chloroquine and hydroxychloroquine groups, n=20 for the control group). Thus, while we demonstrated positive results, the study per se was underpowered. We used a one-month study period and a sample size of 30 (n=18 for (hydroxy)chloroquine vs. n=12 for control) to conduct a post-hoc power analysis. With the alpha level (single-sided) set at 0.025, we would only expect 13.7% (chloroquine vs. control) or 9.2% (hydroxychloroquine vs. control) power, respectively, for detection of a real difference in the median numbers of TTCR days (5.5 days, 6.0 days, and 7.5 days) for chloroquine, hydroxychloroquine and control treatment, respectively.

In this study, the dosage regimen for chloroquine phosphate administration was based on the desire to maximize efficacy while minimizing safety concerns. According to the prescription label of chloroquine phosphate in China, the drug is licensed for the treatment of malaria, extraintestinal amebiasis, and rheumatoid arthritis. When treating acute infectious diseases such as COVID-19, the key to achieve therapeutic efficacy may be finding a path to ensure that blood levels of chloroquine rapidly reach steady state. Because treatment with chloroquine may lead to cardiotoxicity, we excluded patients with prior history of cardiac disorders and prohibited the use of medications (e.g. quinolones) that may lead to QT elongation or ventricular arrhythmia. Therefore, in our study we cautiously used a dosing regimen similar to that used for extraintestinal amebiasis: a 1000 mg QD loading dose of chloroquine phosphate on day 1, followed by a 500 mg QD dose for up to 10 days [12]. Noticeably, the daily and total amounts of chloroquine given in our study were less potent and presumably safer than those recommended dosing regimens by the Chinese Diagnosis and Treatment Protocol for Novel Coronavirus Pneumonia 6^th^/7^th^ editions (500 mg BID for up to 10 days).

At the time when we were preparing this manuscript, there were no *in vitro* data or clinical data on the inhibitory effect of hydroxychloroquine with regard to SARS-CoV-2. However, hydroxychloroquine and chloroquine are highly similar in terms of molecular structure and mechanism of action. Hydroxychloroquine has a better safety profile and is prescribed more commonly. Therefore, when we designed the study, we included hydroxychloroquine for exploratory purposes, without a solid understanding of the optimal dosing regimen. Because of safety concerns, we used the dosage of hydroxychloroquine sulfate for rheumatoid arthritis by the China Food and Drug Administration (200 mg BID for 10 days). Yao et al. recently published the results of *in vitro* experiments showing that the antiviral activity of hydroxychloroquine sulfate against SARS-CoV-2 is greater than that of chloroquine phosphate [19]. In a recently published study of hydroxychloroquine for the treatment of moderate COVID-19, 200 mg BID, 5-day treatment with hydroxychloroquine showed significantly improved TTCR, compared with control treatment [20]. Nevertheless, in our study, hydroxychloroquine treatment did not appear superior to chloroquine treatment. The effect of hydroxychloroquine, compared with control treatment, in shortening TTCR, did not reach the pre-defined criteria for statistical significance. This may be implicated by the following three factors. First, our sample size was small. Second, we used Kaplan-Meier TTCR curves/Logrank testing, which is not a point estimation of efficacy but, rather, a rigorous statistical assessment. Third, the regimen used for dosing hydroxychloroquine may require further optimization. In our study, we used the dose recommended for the treatment of rheumatoid arthritis. This dosing regimen has a good safety profile but may require more time to achieve steady-state levels in the blood. Indeed, in a nonrandomized study of hydroxychloroquine in French patients with different forms of COVID-19, 200 mg TID dosing of hydroxychloroquine for 10 days was significantly associated with viral RNA decline and negativity. Furthermore, these effects were reinforced by combined treatment with azithromycin [21]. However, in another open-label, randomized controlled trial of hydroxychloroquine in mild/moderate or severe COVID-19, Tang et al. demonstrated that hydroxychloroquine did not result in a faster conversion of viral RNA negativity but did alleviate clinical symptoms more effectively than the standard of care alone in patients who did not receive antiviral treatment [22]. In this study, they used a single loading dose of 1,200 mg daily for three days, followed by a maintenance dose of 800 mg daily for the remaining duration of the study period (2 weeks for mild/moderate patients, 3 weeks for severe patients). Taken together, the therapeutic benefit of hydroxychloroquine for COVID-19 may come from its complexed antiviral as well as immunomodulatory effects. While high-dose hydroxychloroquine can suppress the inflammatory response implicated in clinical symptoms, whether it also affects antiviral immunity requires further evaluation. Additional studies will be necessary to optimize patient selection and dosing.

The mode of action underlying these effects remains unclear. Empirical experience and *in vitro* studies have shown that chloroquine may inhibit the production of inflammatory mediators, such as cytokines. However, our results did not support such a mechanism: we observed no significant difference in the levels of inflammatory cytokines before vs. after treatment. The patients enrolled in the study had moderate COVID-19, with limited inflammation. We did not observe changes in patterns of lymphocyte infiltration or SARS-CoV-2-specific IgG/IgM production (cellular and humoral immunity, respectively).

Both chloroquine and hydroxychloroquine were well tolerated and considered safe. Adverse effects were generally mild and comparable between the chloroquine and hydroxychloroquine groups. No serious adverse event occurred. There were modest increases in gastrointestinal symptoms (diarrhea and nausea) in both treatment groups, which can be relieved with symptomatic treatment without affecting subsequent dosing. One patient discontinued chloroquine phosphate on the fifth day of treatment because of elevated ALT/AST levels. The patient’s liver enzyme abnormalities were mild (CTCAE grading) and were not associated with elevated levels of bilirubin. One patient in the hydroxychloroquine group had increased baseline levels of ALT/AST at baseline and ALT/AST levels further increased after dosing, with ALT increase reaching Grade 2 (CTCAE grading). After finishing treatment with hydroxychloroquine, levels of ALT/AST declined. In both patients, the anti-COVID-19 treatments were effective. Our study demonstrated that it is therefore feasible to treat COVID-19 with chloroquine, according to the dosage regimen approved for other indications, as long as the patient’s safety is monitored (e.g. ALT/AST measurement, cardiac monitoring).

Chinese Diagnosis and Treatment Protocol for Novel Coronavirus Pneumonia evolved throughout the study period. During the early days of the study, some patients with pulmonary manifestations, positive SARS-CoV2–specific antibody but negative viral RNA were included. At the later stage of our clinical trial, fewer patients were admitted into the ward, and patient enrollment in the study was discontinued. While our study provides some evidence of the clinical utility of chloroquine, the results need to be further verified by larger trials designed to determine the optimal dosing regimen.

## CONCLUSION

We evaluated the effectiveness of (hydroxyl)chloroquine in treating patients with moderate COVID-19. Our data suggest that chloroquine is likely to be effective in treating certain types of the disease. At the dosage used, hydroxychloroquine exerted effects similar to those of chloroquine; however, the magnitude of hydroxychloroquine’s effects was limited.

## Study highlights

### What is the current knowledge on the topic?

*In vitro* studies have demonstrated that (hydroxyl)chloroquine can inhibit SARS-CoV-2 at the concentrations used for approved indications. However, the translatability of drug’s in vitro activity into efficacy and safety in patients with variable manifestations of COVID-19 will need to be carefully assessed in prospective, randomized and controlled trials.

### What question did this study address?

Whether (hydroxyl)chloroquine, administered at dosages similar to those used for approved indications, affects the clinical recovery of patients with moderate forms of COVID-19.

### What does this study add to our knowledge?

Chloroquine, when used at dosages similar to its approved indication (extraintestinal amebiasis), accelerated recovery from moderate COVID-19. Dosing hydroxychloroquine with the regimen used for rheumatoid arthritis showed a similar but less robust trend. (Hydroxyl)chloroquine were generally safe with acceptable tolerability.

### How might this change clinical pharmacology or translational science?

Modified the dosage regimen used in administering chloroquine to treat amebiasis may be effective for treating certain forms of COVID-19. The dosage regimen for hydroxychloroquine in treating COVID-19 needs further optimization. Our data warrant further investigation of these drugs in large cohort studies.

## Data Availability

The corresponding author are responsible for interpreting all data referred to in the manuscript

## Contributors

Lan Chen, Dan-Yang She, Yong-Hong Zhu, and Zhen-Yu Yin wrote the manuscript; Lan Chen, Dan-Yang She, Yong-Hong Zhu and Zhen-Yu Yin designed the study; Lan Chen, Zhen-Yu Zhang, Zhi-Peng Feng, Su-Zhen Zhang, Qiu-Ying Han, Xiao-Bin Zhang, Xiong Xiao, Hui-Min Chen, Li-Long Liu, Xian-Li Chen, Yu-Pei Lan, De-Jin Zhong, Lan Hu, Jun-Hui Wang, and Xing-Hua Yu conducted the study; Jian-Guo Fu, Lan Chen, Dan-Yang She, Yong-Hong Zhu, and Zhen-Yu Yin analyzed the data; Lan Chen, Dan-Yang She, Yong-Hong Zhu, and Zhen-Yu Yin contributed analytical tools. All authors contributed to data interpretation and manuscript revision. All authors approved the final manuscript.

## Data sharing

We believe that the study plan/procedure, study participants, clinical, immunological, safety and virology data, and statistical analyses were clearly documented in this article. Furthermore, we are willing to share the raw data findings from the study with interested parties (e.g. academics, government, industry, non-profit organizations) upon official request. Participant data will be de-identified except regarding for those pertaining to ethnicity, sex, and age. The robust synopsis will be accessible online following online after publication of this article. Requests for raw data should be submitted in writing to the corresponding authors. Data and the clinical protocol will typically be shared under the conditions of a signed agreement and as otherwise deemed appropriate.

## Acknowledgments

We thank all patients and their families involved infor contributing to the study. We thank Jiancheng Cheng (Shanghai Henlius Biotech, Inc.) for providing statistical advice.

## SUPPLEMENTARY MATERIALS

**Table S1** Baseline disease characteristics of patients with moderate COVID-19 a:Kruskal-Wallis test;b:χ2 test;c:Analysis of Variance (ANOVA)

**Table S2** Key laboratory changes over time

#:ANOVA: P<0.05, compared with the hydroxychloroquine group at Day 7

**Supplementary Materials:** Study Design Protocol

